# Expansion of 5’ UTR CGG repeat in RILPL1 is associated with oculopharyngodistal myopathy

**DOI:** 10.1101/2021.09.18.21263669

**Authors:** Xinzhuang Yang, Dingding Zhang, Pidong Li, Jingwen Niu, Dan Xu, Xueyu Guo, Zhen Wang, Yanhuan Zhao, Haitao Ren, Chao Ling, Yang Wang, Jianxiong Shen, Yicheng Zhu, Depeng Wang, Liying Cui, Lin Chen, Yi Dai

## Abstract

Oculopharyngodistal myopathy is an adult-onset degenerative muscle disorder characterized by ptosis, ophthalmoplegia and weakness of the facial, pharyngeal and limb muscles. Trinucleotide repeat expansions in non-coding regions of LRP12, G1PC1and NOTCH2NLC were recently reported to be the etiologies for OPDM. However, a significant portion of OPDM patients still have unknown genetic causes. In this study, we performed long-read whole-genome sequencing in a large five-generation family of 156 individuals, including 22 patients diagnosed with typical OPDM and identified CGG repeat expansions in *RILPL1* gene in all patients we tested while not in unaffected family members. Methylation analysis indicated that methylation levels of the *RILPL1* gene were unaltered in OPDM patients, which was in consistent with previous reports. Our findings first provided evidences that *RILPL1* were associated OPDM which we suggested as OPDM type 4.

## Introduction

Oculopharyngodistal myopathies (OPDM) are a group of autosomal dominant, adult-onset neuromuscular diseases. The clinical features of OPDM are slowly progressive ptosis, ophthalmoplegia, facial and bulbar weakness and distal predominant limb muscle weakness and atrophy. The disease was first reported and nominated in 1977[1], since then several hundred cases have been reported. The main diagnostic criteria are typical clinical manifestations, inheritance pattern and characteristic muscle biopsy findings including chronic myopathic change with rimmed vacuoles. With the development of long-read sequencing platform, three causative gene locations have been successfully discovered. OPDM type 1(OPDM1; MIM 164310) [2]is caused by the expansion of CGG repeats in the 5’-untranslated region (5’UTR) of *LRP12* gene. OPDM type 2(OPDM2; MIM 618940) [3] is due to the CGG repeat expansion in the 5’UTR of *GIPC1* gene. Meanwhile, the CGG repeat expansion in the 5’UTR of *NOTCH2NLC* gene is responsible for OPDM type 3(OPDM2; MIM 619473)[4]. Many patients with OPDM phenotype could be classified into one of the three types above, but still some patients and pedigrees with identical manifestations were excluded by genetic tests of the CGG repeat expansions in these three loci. Therefore, unknown genetic causes of OPDM, especially expansion of repetitive sequence in other genes, need to be further identified.

The expansion of tandem repeat length causing human disorders almost exclusively affect the neurological system[5]. Autosomal-dominant familial adult myoclonic epilepsy (FAME) characterized by myoclonus and epilepsy is another example[6]. To date, at least the six genes harboring ATTTT/ATTTC expansion leading to FAME are confirmed[7-10]. These six genes have completely different functions and expression profiles. This fact strongly suggests that the pathogenicity of ATTTT/ATTTC repeats is independent of the recipient gene and its function, and that the gene is only a vessel for repeat expression. Similar to FAME, the three genes harboring CGG repeat expansions in 5’UTR have totally distinct functions and expression patterns. The CGG repeat expansion in *NOTCH2NLC* was initially recognized as the genetic cause for neuronal intranuclear hyaline inclusion disease and other neurodegenerative diseases affecting the central nervous system[11-15]. Based on this knowledge, we hypothesize that the unidentified pathogenicity of OPDM could be the CGG repeat expansion located in the untranslated region of a new gene.

In this study, we explored other gene loci for repeat expansions in a large Chinese pedigree with autosomal dominant OPDM. Firstly, the patients in this family were excluded OPMD and OPDM type 1, 2, and 3 by Repeat-Primed PCR. By using long-read whole-genome sequencing (LRS) on the Oxford Nanopore platform and PacBio SMRT platform, we found a CGG repeat expansion in the 5’UTR segment of *RILPL1* gene, which had not been related to any monogenic disease, was associated with OPDM. We suggest that the new *RILPL1*-defined disease entity could be designated as OPDM type 4 (OPDM4).

## Result

### Clinical and pathological features

A large five-generation Chinese family of 156 individuals, including 22 patients diagnosed with OPDM were recruited (Fig. 1). Patients with OPDM showed slowly progressive weakness in extraocular, laryngopharyngeal, facial, and upper and lower limb muscles. The weakness of limbs was distal predominance. In this family, the onset symptom was insidious, which was uniformly ptosis and ophthalmoparesis. The patients gradually developed dysphagia, dysarthria and limb weakness in the following years, and in the end stage, became bed-ridden. The intrafamily heterogeneity was obvious. Some patients remained stable in decades, while others experienced apparent progression in a few years. We followed up the index patient for more than 3 years. His main symptoms were limited to external ocular muscles with a course longer than 10 years. But the MR scan of his thigh muscles revealed a subclinical progressive fat infiltration in inner thigh muscles during a 3-year period. (Fig. 2) The proband had complaints of palpitation and chest distress. Therefore, we performed gadolinium-enhanced cardiac MR imaging. We found delayed myocardial enhancement with a normal heart size and function. To the best of our knowledge, this is the first report of myocardial impairment in OPDM (Fig. 2)

**Figure 1.**
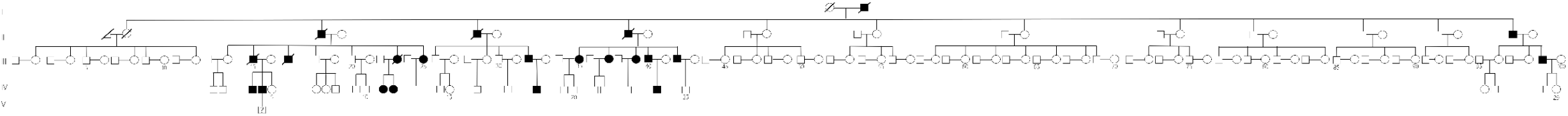
Pedigrees for the Chinese OPDM family presented in this study

**Figure 2.**
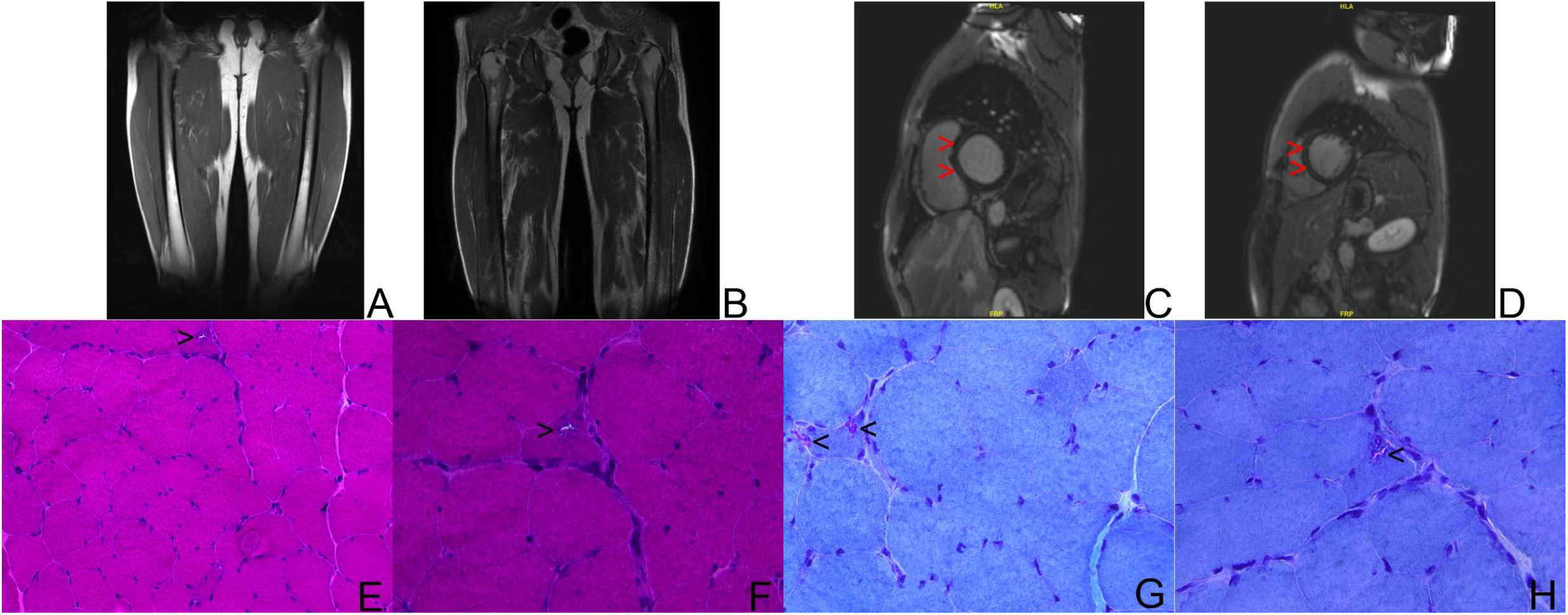
MR image and pathology of patient III-42. (A–B) T1-weighted MR images of thigh of index patient. A. image obtained three years ago; B. image obtained this year. Fatty infiltration in inner muscles was obvious this year, compared with almost no lesion in thigh muscles three year ago. (C-D) Gadolinium-enhanced cardiac MR imaging. Delayed myocardial enhancement was found in the middle of left ventricular wall (red arrow head). (E-H) Skeletal muscle pathology in OPDM. (E-F) Haematoxylin and eosin staining showed mild variation in fiber size and rimmed vacuoles. (black arrow head), E. Magnification X20; F. magnification X20. (G-H) Modified Gomori Trichome staining demonstrated rimmed vacuoles. (black arrow head) G-H magnification X40

The pathological findings of muscle biopsy were in accordance with classical OPDM features. The histopathology of the quadriceps muscle showed mild myopathic changes with increase in fiber size and rimmed vacuoles in a few muscle cells (Fig. 2)

### Identification of CGG repeat expansions in *RILPL1* gene in patients with OPDM

To determine the genetic causes for patients in this family, we first checked the repeat expansion in *PABPN1, LRP12, G1PC1* and *NOTCH2NLC* which are pathogenic genes for OPDM type 1-3 and OPMD. However, they were all excluded by RP-PCR (Supplementary Figs.1A-1H), indicating that there was a novel genetic mechanism in this large OPDM family. Subsequently, we performed whole-exome sequencing of five affected and five unaffected members from this family, while the single nucleotide variant and copy number variant analysis did not reveal potential associated events (data not shown).

Considering the pathogenic mechanism for OPDM are all repeat expansions on genes’ 5’UTR according to previous reports, we performed long-read whole-genome sequencing by ONT PromethION sequencing platform on 4 affected (III-37, III-39, III-40 and III-42) and 3 unaffected family members (II-8, III-41 and IV-21)[16]. Gene coverage reached 98.73% in average, and mean depth is about 14.8X. Taking the advantage of long-read sequencing, repeats expansion exploration was performed by our previously reported STR-scoring method, which was a strategy to identify expanded repeats in the long reads based on comparisons between healthy and affected individuals (Material and Methods)[3]. As results, 107, 287 short tandem repeats were identified in whole genome. Among the top10 candidate repeats, a heterozygous CGG repeat expansion in the upstream of *RILPL1* have a considerable high score (chr12:124, 018, 268-124, 018, 302, Fig. 3A, and Supplementary Table 1). To avoid potential drawbacks of the pipeline which gave different weight factors to different gene regions and different patterns to the score, we re-evaluated the scores of those STRs without weight coefficients and confirmed that the CGG repeats in the upstream of *RILPL1* was still the most significant locus (Supplementary Table 2). The repeat counts of this locus in 4 patients in this family were 107∼187, while only 31∼33 in heathy individuals (Fig. 3B, 3C). To further confirm the result of ONT, we performed PacBio sequencing for patients III-39, which identified expanded GGC repeats in the same locus of *RILPL1* as with ONT data (Supplementary Fig. 2). All the above provided the evidence for the association between the CGG repeat expansion in *RILPL1* and OPDM in our study. Considering RNA toxicity is a common mechanism behind many human neurological diseases and disorders caused by abnormal non-coding trinucleotide repeat expansions[2, 5, 16], we further examined whether it was the case for OPDM. According to gene structures annotated by RefSeq, Ensembl and GENCODE, the CGG repeat is located 3bp upstream of *RILPL1* gene (Material and Methods, Supplementary Fig. 3A). However, three independent pieces of evidence suggested the existence of an alternative transcription start site (TSS) upstream of CGG repeat. First, two RNA ESTs, including one spliced one, started from approximately 230bp upstream of *RILPL1* (Supplementary Fig. 3A). Second, consistent with EST evidence, CAGE-seq of skeletal muscles from FANCOM5 database also suggested an alternative TSS (Material and Methods, Supplementary Fig. 3A). Third, by analyzing our in-house RNA-seq data of skeletal muscles, we noted substantial RNA-seq signals upstream of the annotated TSS (Material and Methods, Supplementary Fig. 3A). In sum, we concluded that the transcription of *RILPL1* might initiate from an upstream TSS and include CGG repeats as part of its 5’UTR. This suggested that RNA toxicity was a plausible mechanism for OPDM that awaited future investigation.

**Figure 3.**
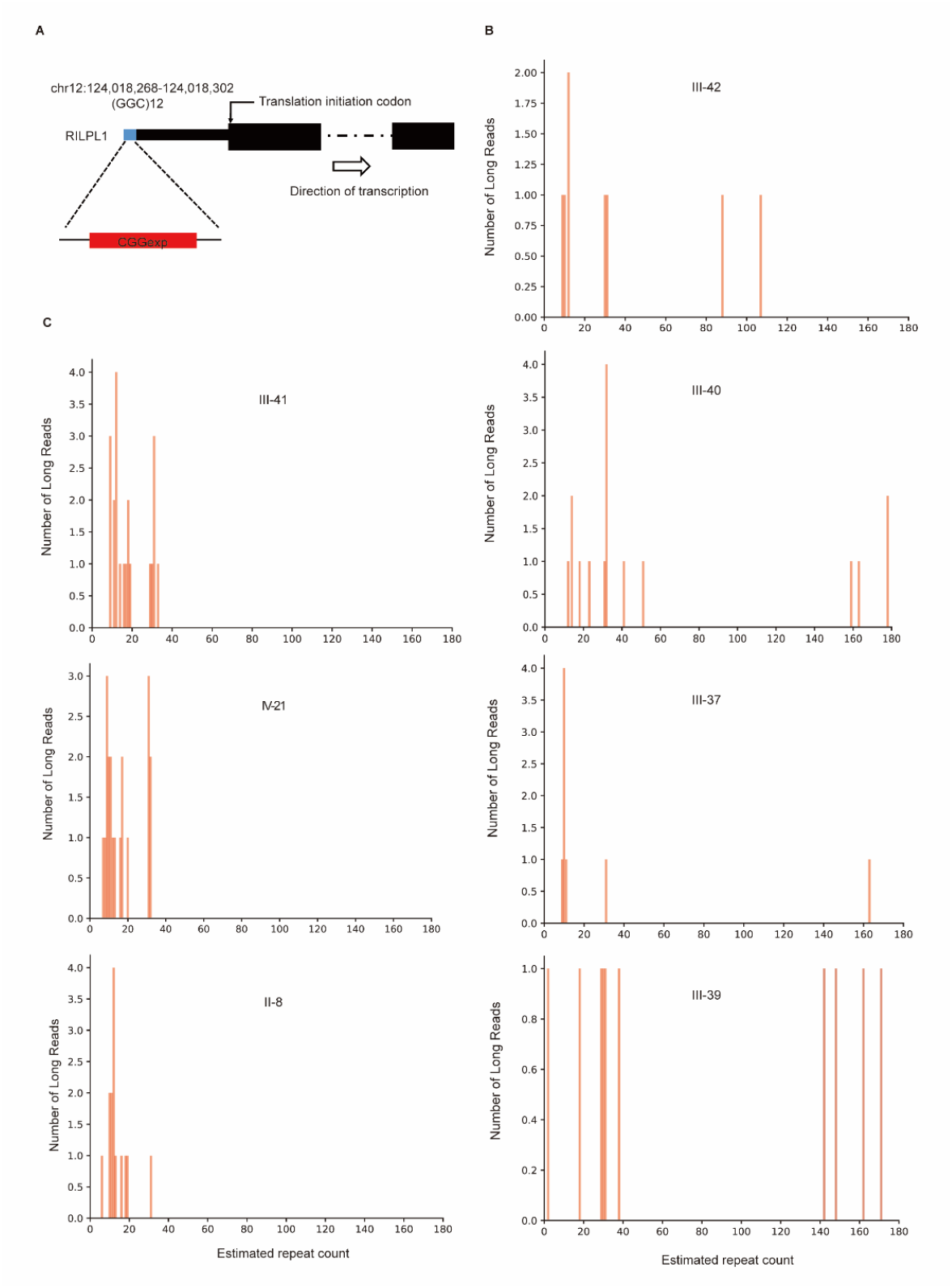
The genomic position and repeat counts of the CGG expansion. (A) Genomic position of the CGG repeat expansion in RILPL1. Estimated repeat counts of the CGG expansion were more than 100 in four patients (III-37, III-39, III-40 and III-42) (B) and only 30 in three healthy family members (II-8, III-41 and IV-21) (C), as analyzed by long-read whole-genome ONT sequencing platform.

### Methylation Status and RNA level of *RILPL1* in OPMD4

To further evaluate the underlying mechanism of CGG repeat expansions, we performed 5mc (5-methylcytosine) DNA modification surrounding CGG repeats and adjacent CpG islands with ONT PromethION sequencing data (Material and Methods). Consist with other OPDM repeat-associated pathogenesis, there was no difference in DNA methylation levels between patients and healthy individuals (Wilcoxon test, P-value=0.30, Fig. 4A)[3]. Interestingly, even no DNA methylation signal was detected on both groups. As positive control, we found increased 5mc methylation level in Alu repeat sequence regions (Fig. 4B). To further examine whether RNA level for *RILPL1* has changed within patients, we conducted RNA-sequencing with muscle tissues to analyze the transcription level. The results presented no significant difference between patient III-42 and age-matched controls (Material and Methods, Fig. 4C, supplementary Table. 3).

**Figure 4.**
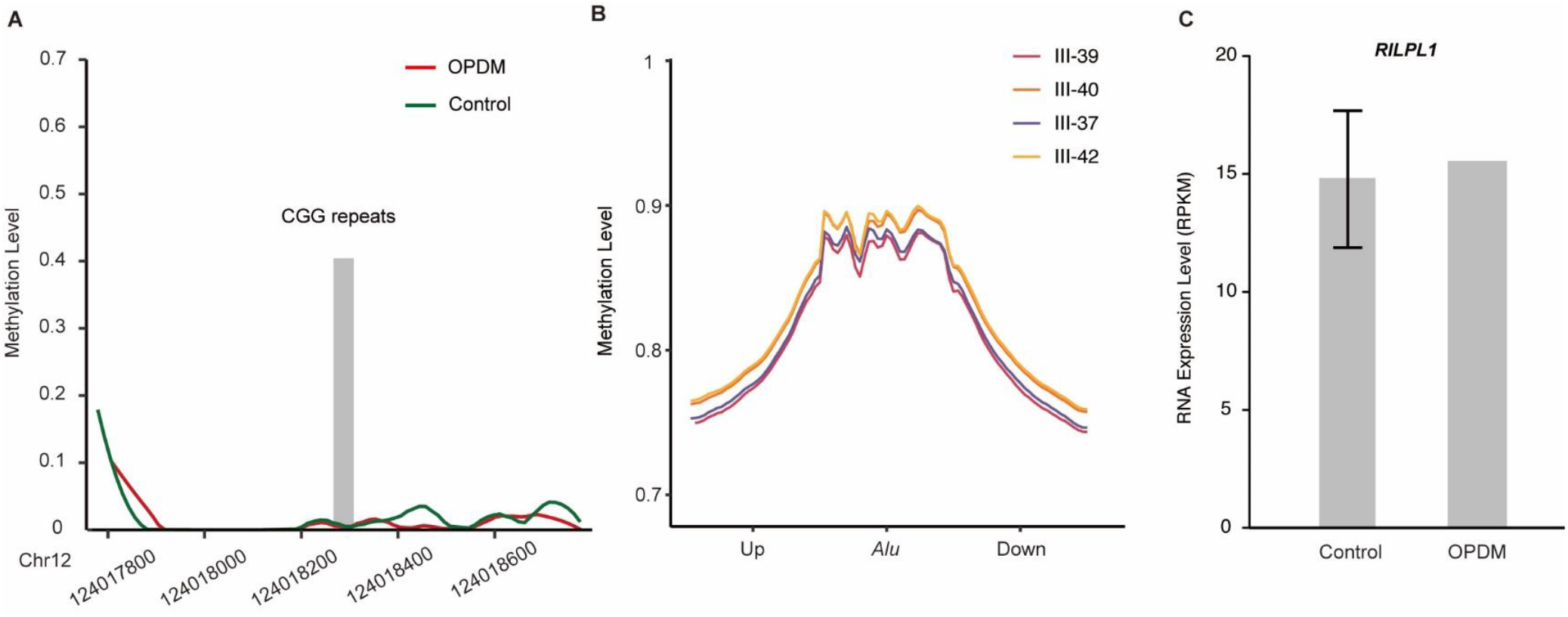
Methylation and RNA Expression of *RILPL1* gene (A) Methylation status across the expanded GGC repeat region in *RILPL1* was determined using ONT data from four affected (III-37, III-39, III-40 and III-42) and three unaffected family members (II-8, III-41 and IV-21); no significant difference in methylation was detected between two groups. (B) Methylation status in Alu sequences along whole human genome, which are generally highly methylated in four RILPL1-affected individuals with OPDM, as positive controls. (C) RNA-seq analysis of *RILPL1* RNA expression levels in patient III-42 and three age-matched controls; no significant different was detected.

**Table 1.**
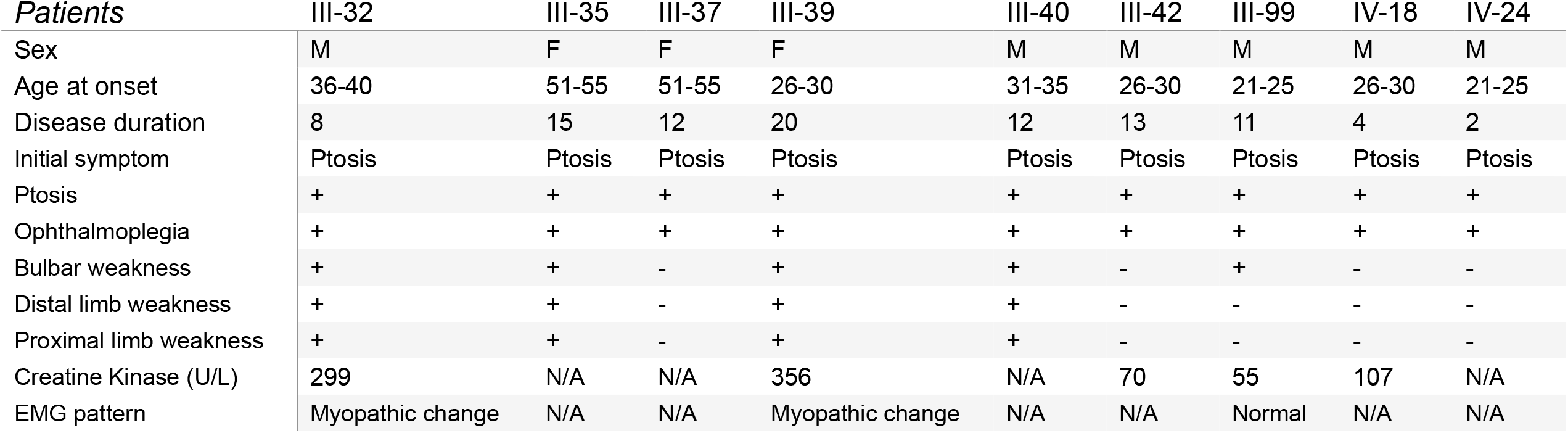
Clinical features of affected individuals in the pedigree

## Material and Methods

### Library preparation and Whole-exome Sequencing

The exome sequences were efficiently enriched from 0.4 μg genomic DNA using Agilent liquid capture system (Agilent SureSelect Human All Exon V6) according to the manufacturer’s protocol. DNA libraries were sequenced on Illumina NovaSeq for paired-end 150bp reads. Valid sequencing data was mapped to the reference genome (GRCh37/hg19) by Burrows-Wheeler Aligner (BWA) software to get the original mapping results in BAM format. Subsequently, GATK (Version) were used to do variant calling and identify SNP and indels. Then, Conifer (Version) were used to identify CNVs. At last, ANNOVAR was performed to do annotation for VCF (Variant Call Format) file.

### Long-Read Whole-Genome Sequencing

DNA samples of affected individuals with OPDM and healthy individuals were sequenced using PromethION sequencer (Oxford Nanopore Technologies). Library preparation was carried out using a 1D Genomic DNA ligation kit (SQKLSK109) according to the manufacturer’s protocol. For each individual, one PRO-002 (R9.4.1) flowcell was used. PromethION data base-calling was performed using guppy v.3.3.0 (Oxford Nanopore Technologies), and only pass reads (qscore≥ 7) were usedfor subsequent analysis. Long reads were aligned to reference genome (GRCh37/hg19) by minimap2 (v2.8, Li, H. (2018). Minimap2: pairwise alignment for nucleotide sequences. Bioinformatics 34, 3094–3100.). To validate the accuracy of ONT, we also performed PacBio Single Molecule, Real-Time (SMRT) DNA sequencing for patients FIII-39. HQRF (High quality Region Finder) were used to identify the longest region of singly-loaded enzyme activity. low-quality areas were filtered by Signal Noise Ratio (SNR). Subreads were obtained after the basic filtrations as previous reported. Circular Consensus Sequence (CCS) reads were retained using CCS tools and then aligned to the reference genome (GRCh37/hg19) using PBmm2.

### STR Detection and STR-Scoring Framework

The short tandem repeats that located in genic regions including 10 Kb up-/downstream of genes were identified in whole genome, using long-read whole-genome sequencing based on the STR-Scoring framework, as previously described (Expansion of GGC Repeat in GIPC1 Is Associated with Oculopharyngodistal Myopathy). Considering that nanopore reads have certain bias errors, we used IGV software (Helga Thorvaldsdóttir, James T. Robinson, Jill P. Mesirov. (2013). Integrative Genomics Viewer (IGV): high-performance genomics data visualization and exploration. Briefings in Bioinformatics 14, 178-192) to further confirm and correct the repeat count of each read for the top 10 STRs, and refined the second largest repeat count as the estimated repeat count (ERC).

### 5mc DNA Methylation analysis

The Minimap2 (v2.8, Li, H. (2018). Minimap2: pairwise alignment for nucleotide sequences. Bioinformatics 34, 3094–3100.) and Nanopolish (v0.9, Simpson, J.T., Workman, R.E., Zuzarte, P.C., David, M., Dursi, L.J., and Timp, W. (2017). Detecting DNA cytosine methylation using nanopore sequencing. Nat. Methods 14, 407–410.) were applied for 5mC DNA methylation calling in ONT sequencing data. The methylation level around the CGG repeats and adjacent CpG island was compared between affected individuals and healthy individuals, and between expanded and non-expanded alleles using the Wilcoxon Rank Sum Test. Adjacent GpG was defined as chr12: 124,017,594-124,018,994, and CGG repeat region were defined as chr12:124,018,268-124,018,302 (GRCh37/hg19).

### RNA-seq Analysis

Total RNAs were extracted from muscle tissues of patients III-42 and three controls following Trizol RNA isolation procedure (Supplementary Table. 3). The quality of the input RNA was controlled using Agilent 2100. Total RNA samples were then applied to strand-specific, poly(A)-positive RNA-seq following the manufacturer’s protocols and pipelines as previously described[17, 18]. Deep sequencing was then performed on Illumina Hiseq X Ten sequencing systems with 151-bp paired-end reads mode. RNA-seq data were then aligned to human genome (GRCh37/hg19) by HISAT2 (V2.1.0) and read count were calculated by HTseq (V0.11.2). Gene expression level measured by RPKM were performed by our in-house Perl scripts[17].

### Identification of novel transcriptional start site of RILPL1

EST (Expressed Sequence Tag) data were obtained from UCSC (http://genome.ucsc.edu/,GRCh37/hg19) and CAGE-seq (Cap Analysis of Gene Expression AND deep Sequencing, BAM format) were download from FANTOM5 (https://fantom.gsc.riken.jp/5/) by searching for human skeletal muscle tissue data and we finally got three datasets with high data quality. BAM files were transformed into bigwig format and uploaded on UCSC browser for visualization.

### Repeat-primed PCR (RP-PCR)

Genomic DNA of patient III-42 from this family was analyzed by repeat-primed PCR (RP-PCR). The PCR mix contained 0.25 U PrimeSTAR GXL DNA Polymerase, 13 PrimeSTAR GXL Buffer, 200 mM each dATP, dTTP, dCTP (Takara Bio, Shiga), and 7-deaza-dGTP (Sigma-Aldrich), 5% dimethyl sulfoxide (SigmaAldrich), 1M betaine (Sigma-Aldrich), 0.3 mM primer F and inker-R, 0.1 mM primer R, and 100 ng genomic DNA in a total reaction volume of 20 mL. Four pair of primers were listed in Supplementary Table. 4. We used a slow-down PCR protocol [19]: initial denaturation at 95 °C for 5 min, followed by 50 cycles of 95 °C for 30 s, 98 °C for 10 s, 62 °C for 30 s and 72 °C for 2 min. The ramp rate to 95 °C and 72 °C was set to 2.5 °C s−1 and that to 62 °C was set to 1.5 °C s−1.Electrophoresis was performed on a 3500xl Genetic analyzer (Thermo Fisher Scientific) and the data were analyzed using GeneMapper software (Thermo Fisher Scientific).

### Ethics issues declaration

The present study has been approved by the Ethics Committees of Peking Union Medical College Hospital. And the written informed consent forms were obtained from all the study participant. Furtherly, we have shared the study findings with all the study participants.

## Discussion

Oculopharyngodistal myopathy (OPDM) is defined as an adult-onset, slowly progressive, autosomal dominant muscle disease. Recently, three different gene loci were identified as causative pathogenesis for OPDM. The phenotypes of OPDM 1, 2 and 3 are highly homogeneous[1], while the genotypes share obvious similarity. All the expansions are CGG repeats located in 5’UTR of the genes. When we found this huge pedigree consistent with all the aspects of OPDM and excluded all the three established OPDM subtypes, we realized that there must be a new repeat expansion located in an unidentified pathogenic gene.

Although traditional genetic linkage analysis could be in used in mapping the rough location of causative gene in this pedigree. With the development of long-read sequencing platform and advanced bioinformatic methods, we directly found a significant signal of a CGG repeat expansion in the upstream region of *RILPL1* using the STR-Scoring pipeline[3]. This expansion presented clear co-segregation in the family. The copy number of CGG repeats in all examined patients was more than 100, while was about 30 in unaffected family members and other healthy individuals. In order to verify the accuracy of ONT reads in identifying CGG repeats, we conducted PacBio sequencing for sample III-39 with higher accuracy, and the results were in consistent with ONT. This is the first time that *RILPL1* gene has been associated with a monogenic disease, OPDM. We call this type of OPDM associated with *RILPL1* OPDM4. We are trying to set up RP-PCR as a simple and low-cost method to detect other members in this large family and other patients and pedigrees.

RILPL1, Rab Interacting Lysosomal Protein Like 1, which was highly expressed in muscle tissues (Supplementary Fig. 4), plays a role in the regulation of cell shape and cellular protein transport. It is also a neuroprotective protein, involved in the regulation of neuron death by sequestering GAPDH in the cytosol and preventing the apoptotic function of GAPDH in the nucleus. RILPL1 was reported to be associated with several neurological diseases, such as epilepsy and familial temporal lobe dementia, while not any description with OPMD.

Previous gene annotations showed that the CGG repeats were located in the upstream region of the transcription start site of *RILPL1* gene, which was nearly 3bp away from TSS. However, our analysis revealed that there was a novel TSS in the upstream region of *RILPL1*. We first found that there were ESTs, supporting the existence of RNA expression beyond the traditional transcription start site. To further confirm this was a novel TSS, we downloaded CAGE data of skeletal muscle tissues from FANTOM5 database, and found that *RILPL1* had another significant CAGE peak in front of the annotated TSS, suggesting the existence of a new TSS. In addition, our analysis of in-house skeletal muscle RNA-seq data also confirmed the existence of RNA expression in the upstream of traditional TSS. Therefore, we proposed that there was a novel TSS and the CGG repeats were located in the 5’UTR of *RILPL1* gene, playing a role in regulating gene transcription, just like the CGG repeat expansions in other types of OPMD.

In order to explore the biological effects of CGG repeat expansions, we first analyzed the DNA methylation level of *RILPL1*. No significant differences in methylation were found between patients and healthy members, which was in consistent with previous reports. We also conducted methylation analysis for two alleles of heterozygotes and found no significant difference between them. It might suggest that CGG amplification may not work on methylation level. We subsequently explored gene expression differences with muscle RNA-seq data among patients and normal controls, and still found no significance on *RILPL1* expression between the two groups. We are now investigating the expression and subcellular localization of *RILPL1* at the protein level and trying to figure out the underlying molecular mechanisms of OPDM4.

All the patients in this family followed the same pattern of disease progression. The onset symptom was exclusively ptosis and then other extraocular muscles, bulbar, distal and proximal limb muscles were sequentially affected. But the progression rate was apparently different. Some patients developed severe limb weakness in less than five years while others only had weakness and atrophy of ocular and pharyngolaryngeal muscle. Interestingly, there was no apparent correlation between the age of onset and the progression rate of the disease. The enhanced MR scan of index patient’s heart showed delayed myocardial enhancement in the left ventricular wall. This phenomenon pointed out that the myocardium was involved in OPDM.

In summary, we found that a CGG repeat expansion in RILPL1 was responsible for OPDM in a large Chinese family. Our study widens the causative genes and loci spectrum of Oculopharyngodistal myopathy.

## Data Availability

we, all the authors, have the availability of all data used in the present manuscript.

## Figure legend

Supplementary Figure 1

The results of Sanger sequencing, PCR Capillary Electrophoresis, and Repeat-Primed Polymerase Chain Reaction Capillary Electrophoresis of patient III-39. No repeat expansions in *PABPN1, LRP12, GIPC1* and *NOTCH2NLC* were detected.

Supplementary Figure 2

The CGG repeat expansion in the 5’UTR of *RILPL1* identified by PacBio sequencing data was consist with ONT.

Supplementary Figure 3

Gene annotation track from NCBI, UCSC, and GENCODE present traditional gene structures of *RILPL1*. While RNA ESTs, RNA-seq, and CAGE-seq provided evidences that *RIPL1* gene transcription might initiate from an upstream TSS and include expanded CGG repeats as part of its 5’UTR.

Supplementary Figure 4

RNA expression level from Consensus dataset, HPA dataset, and GTEx dataset show that *RILPL1* is highly expressed in muscle and brain tissues.

