## Supplementary figures and images for "Expansion of 5’ UTR CGG repeat in RILPL1 is associated with oculopharyngodistal myopathy"

### Supplymentary Figure 1

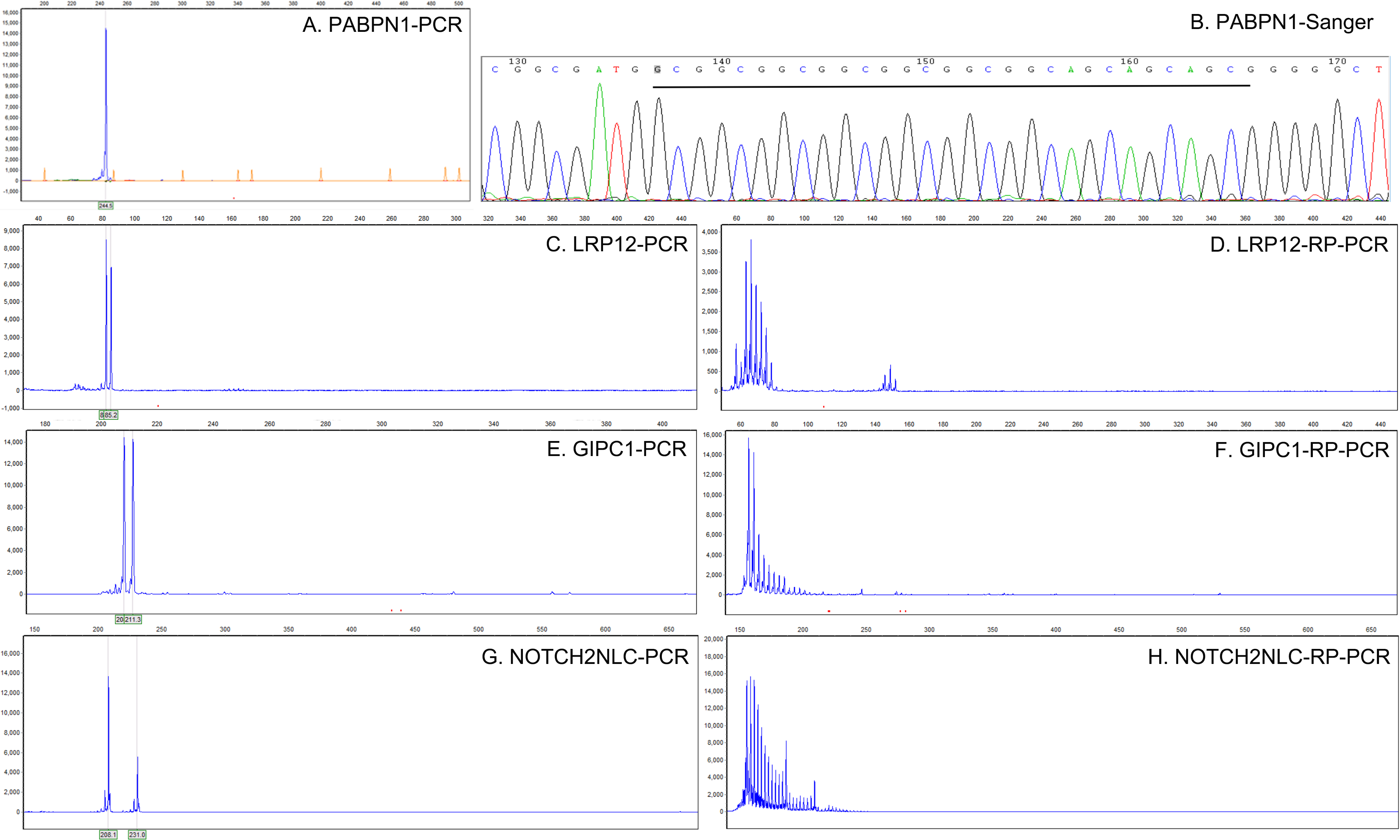

### Supplymentary Figure 2

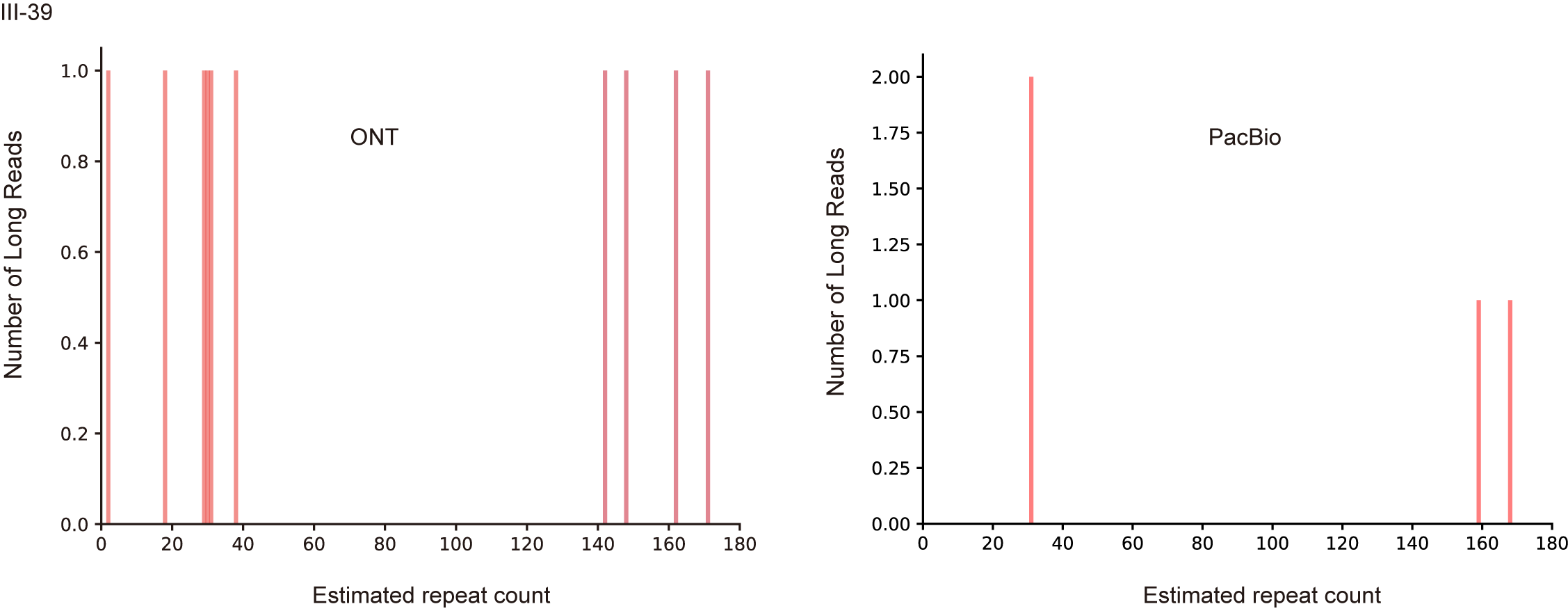

### Supplymentary Figure 3

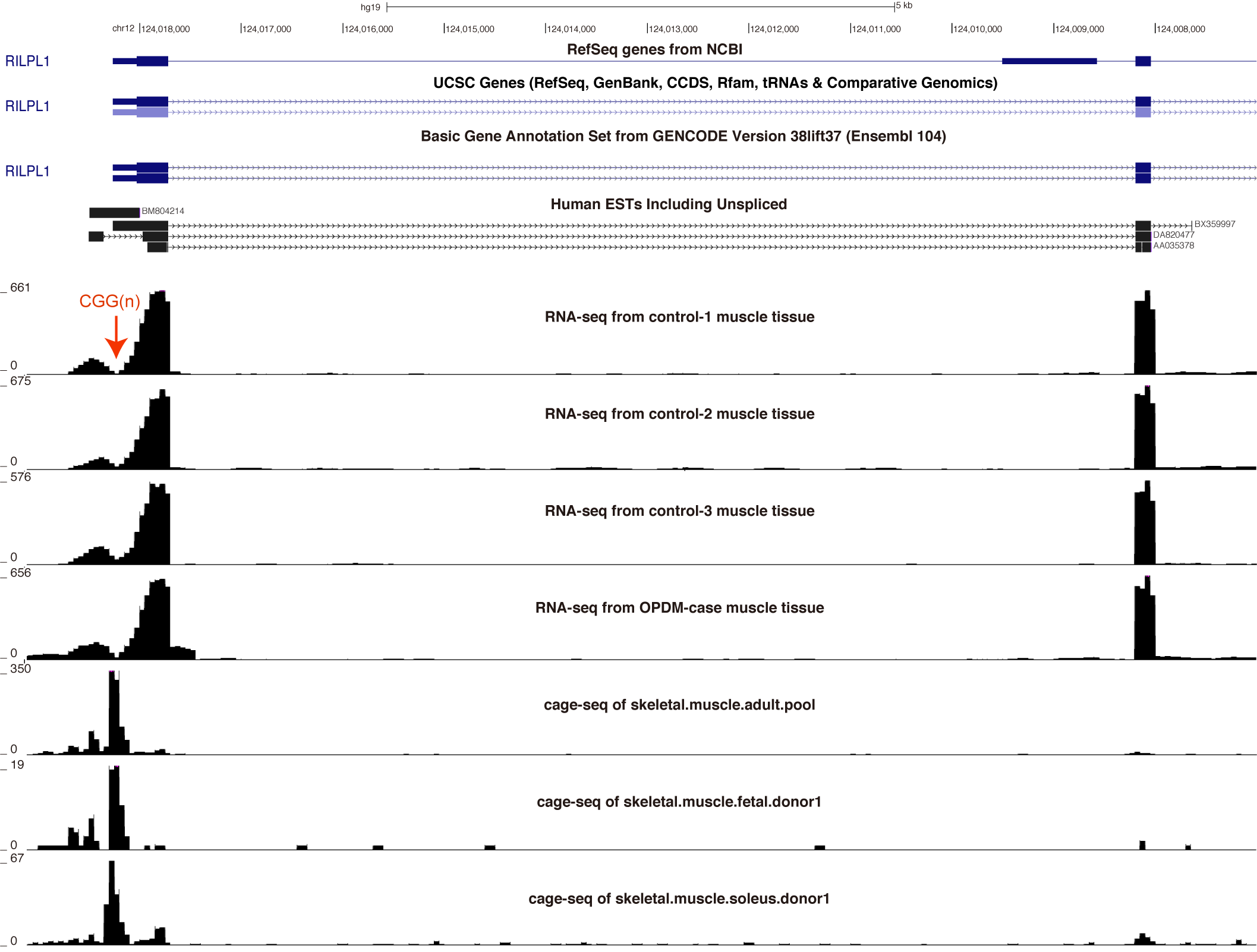

### Supplymentary Figure 4

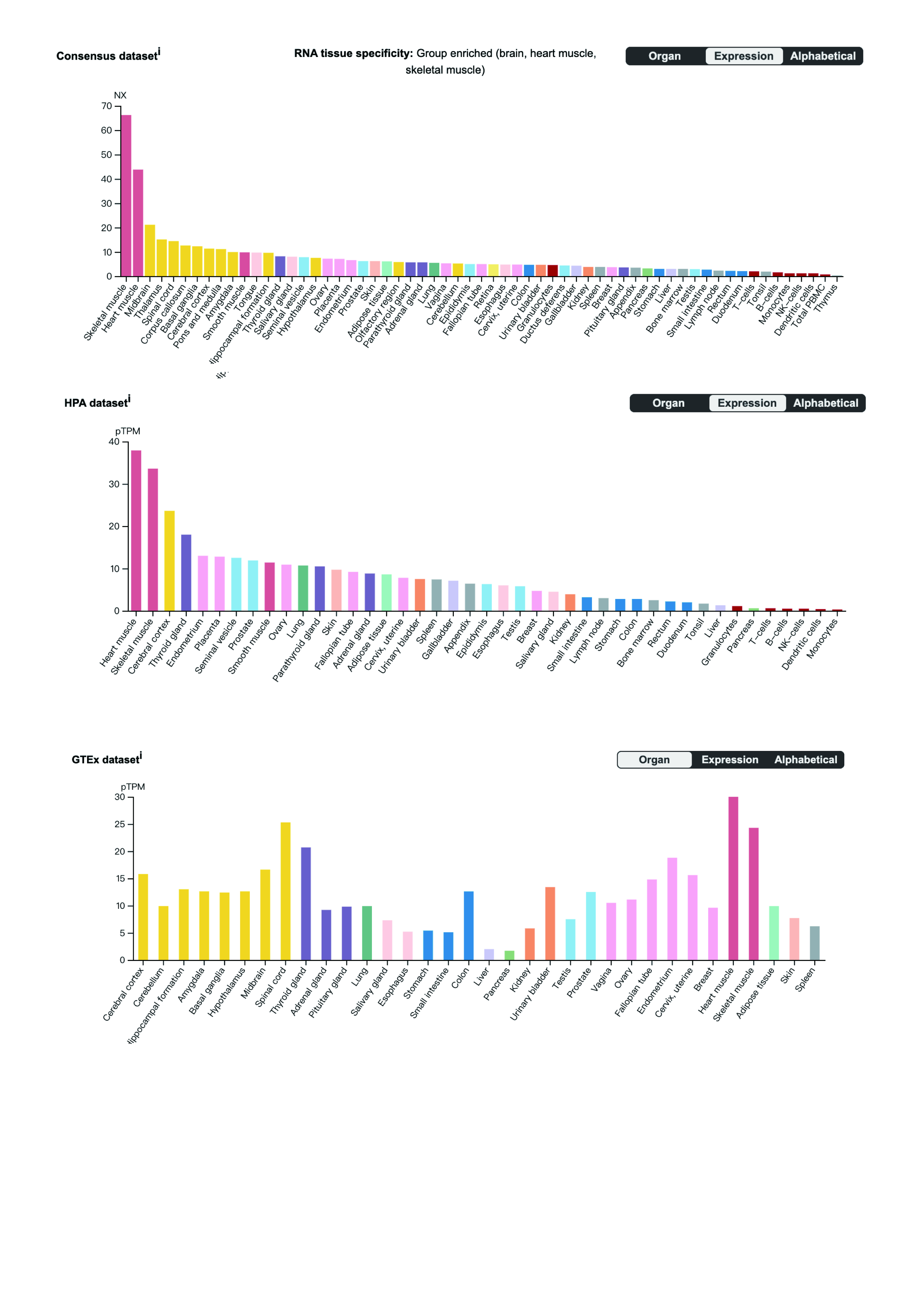
