## Supplementary material for "Expansion of 5’ UTR CGG repeat in RILPL1 is associated with oculopharyngodistal myopathy": Supplymentary Table 3

Supplementary Table. 3. The characteristics of controls who contributed muscle tissue

| Patients | Control-1 | Control-2 | Control-3 |
| --- | --- | --- | --- |
| Sex | F | F | F |
| Age (years) | 61-65 | 66-70 | 66-70 |
| Disease for surgery | Lumbar Spinal Stenosis | Lumbar Spinal Stenosis | Lumbar Spinal Stenosis |
