## Supplementary material for "Expansion of 5’ UTR CGG repeat in RILPL1 is associated with oculopharyngodistal myopathy": Supplymentary Table 4

| gene | direction | Primer |
| --- | --- | --- |
| PABPN1 | forward | CGCAGTGCCCCGCCTTAGA |
|  | reverse | ACAAGATGGCGCCGCCGCCCCGGC |
| GIPC1 | forward | CACATCCTTCTCGCAGAGGCCAC |
|  | reverse | GAAGACGCGGATTGGCTGCGAGC |
| LRP12 | forward | GGGAGGAGAAGCTGGAGGTA |
|  | reverse | GGAGAGCGCAGGGAGCAG |
| NOTCH2NLC | forward | GGCATTTGCGCCTGTGCTTCGGACCGT |
|  | reverse | TACGCATCCCAGTTTGAGACGTCCTCCGCCGCCGCCGCC |

Supplementary Table. 4. Primer sequences for PCR
